# Prediction of type 1 diabetes at birth: cord blood metabolites versus genetic risk score in the MoBa cohort

**DOI:** 10.1101/2021.02.20.21252128

**Authors:** German Tapia, Tommi Suvitaival, Linda Ahonen, Nicolai A. Lund-Blix, Pål R. Njølstad, Geir Joner, Torild Skrivarhaug, Cristina Legido-Quigley, Ketil Størdal, Lars C. Stene

## Abstract

**Background and aim:** Genetic markers are established as predictive of type 1 diabetes, but unknown early life environment is believed to be involved. Umbilical cord blood may reflect perinatal metabolism and exposures. We studied whether selected polar metabolites in cord blood contribute to prediction of type 1 diabetes.

**Methods:** Using a targeted UHPLC-QQQ-MS platform, we quantified 27 low molecular weight metabolites (including amino acids, small organic acids and bile acids) in 166 children, who later developed type 1 diabetes, and 177 random control children in the Norwegian Mother, Father and Child (MoBa) cohort. We analysed the data using logistic regression (estimating odds ratios per standard deviation [aOR]), area under the receiver operating characteristic curve (AUC) and k-means clustering. Metabolites were compared to a genetic risk score based on 51 established non-HLA SNPs, and a four-category HLA risk group.

**Results:** The strongest associations for metabolites were aminoadipic acid (aOR=1.23,95%CI:0.97–1.55), indoxyl sulfate (aOR=1.15,95%CI:0.87–1.51), and tryptophan (aOR=0.84,95%CI:0.65–1.10), with other aORs close to 1.0, and none significantly associated with type 1 diabetes. K-means clustering identified six clusters, none of which were associated with type 1 diabetes. Cross-validated AUC showed no predictive value of metabolites (AUC 0.49), while the non-HLA genetic risk score AUC was 0.56 and the HLA risk group AUC was 0.78.

**Conclusions:** In this large study, we found no support of a predictive role of cord blood concentrations of selected bile acids and other small polar metabolites in the development of type 1 diabetes.

**Tweet:** Predicting childhood type 1 diabetes with cord blood biomarkers: genetic risk score works but metabolites do not

@OsloDiabetes #T1D

## Introduction

Type 1 diabetes is usually preceded by a prodromal phase characterized by islet autoantibodies, often appearing in early childhood years prior to diagnosis. [1] HLA and other genetic factors clearly contribute to type 1 diabetes susceptibility, [1] but the increasing incidence implicate non-genetic factors. [2] The typically early seroconversion to islet autoantibodies suggests that early life is important. [3-5]

Maternal age, obesity and birth weight are early life non-genetic risk factors relatively consistently associated with childhood-onset type 1 diabetes. [6-11] Obesity, dysglycaemia, kidney function and related traits, both in non-pregnant adults and in pregnant women, are associated with perturbations in small metabolites such as amino acids, creatinine, and bile acids, [12-15] many of which have also been associated with birth weight. [12] Metabolites such as glucose, lipids, amino acids and bile acids can cross the placenta, often bidirectionally, via free diffusion and placentally expressed transmembrane transporters. [16-18] Many small metabolites in cord blood are thus correlated with maternal levels during the third trimester. [12] For example, plasma creatinine, a marker of kidney function, largely reflects maternal levels when measured in cord blood [19], but has been linked to birth weight. [20, 21] Maternal circulating bile acids, which may be influenced by maternal gut microbiota, may program offspring metabolism, or influence their microbiome, and have been linked to insulin resistance. [14, 22, 23] Yet, there is only one small study (15 cases and 24 controls) in cord blood, [24] and one study using dried blood spots, [25] to date on non-lipid metabolites and later type 1 diabetes. The authors are not aware of any previous study investigating maternal or new-born plasma bile acids and subsequent type 1 diabetes risk.

The aim of the study was to test if selected small metabolites in cord blood, or combinations of these, could predict future risk of offspring type 1 diabetes in the Norwegian Mother, Father and Child Cohort Study (MoBa), one of the largest pregnancy cohorts in the world. To ensure robust quantification, and to minimise multiple testing problems, we chose a targeted metabolomics approach focusing on small metabolites (molar mass ranging from 75 to ∼500 g/mol) with previous evidence for association with metabolic traits. [13] In addition, we investigated established genetic susceptibility markers for comparison of predictive values among biomarkers present at birth.

## Methods

### Participants and study design

We designed a nested case-control study in the Norwegian Mother, Father and Child Cohort Study (MoBa) [26], which recruited ∼114,000 pregnant mothers (41% eligible participated) nationwide from 1999-2008 (last birth in 2009). The current study uses data from cord blood samples and repeated questionnaires collected during pregnancy and up to child age 6 months [27]. All participating mothers gave written informed consent. The establishment of MoBa and initial data collection was based on a license from the Norwegian Data Protection Agency and approval from The Regional Committees for Medical and Health Research Ethics. The MoBa cohort is now based on regulations related to the Norwegian Health Registry Act. The current study was approved by The Regional Committees for Medical and Health Research Ethics. Children who developed type 1 diabetes by February 5, 2014 were identified by register linkage to the Norwegian Childhood Diabetes Registry, [28] and selected as cases. A random sample of the cohort was included as controls. Case and control samples were retrieved simultaneously and treated equally. In total, 166 children were type 1 diabetes cases, and 177 children were controls (Figure 1). Baseline characteristics for those with available blood samples were largely similar to the whole MoBa cohort, except a lower proportion of caesarean section and preterm birth. [29]

**Figure 1:**
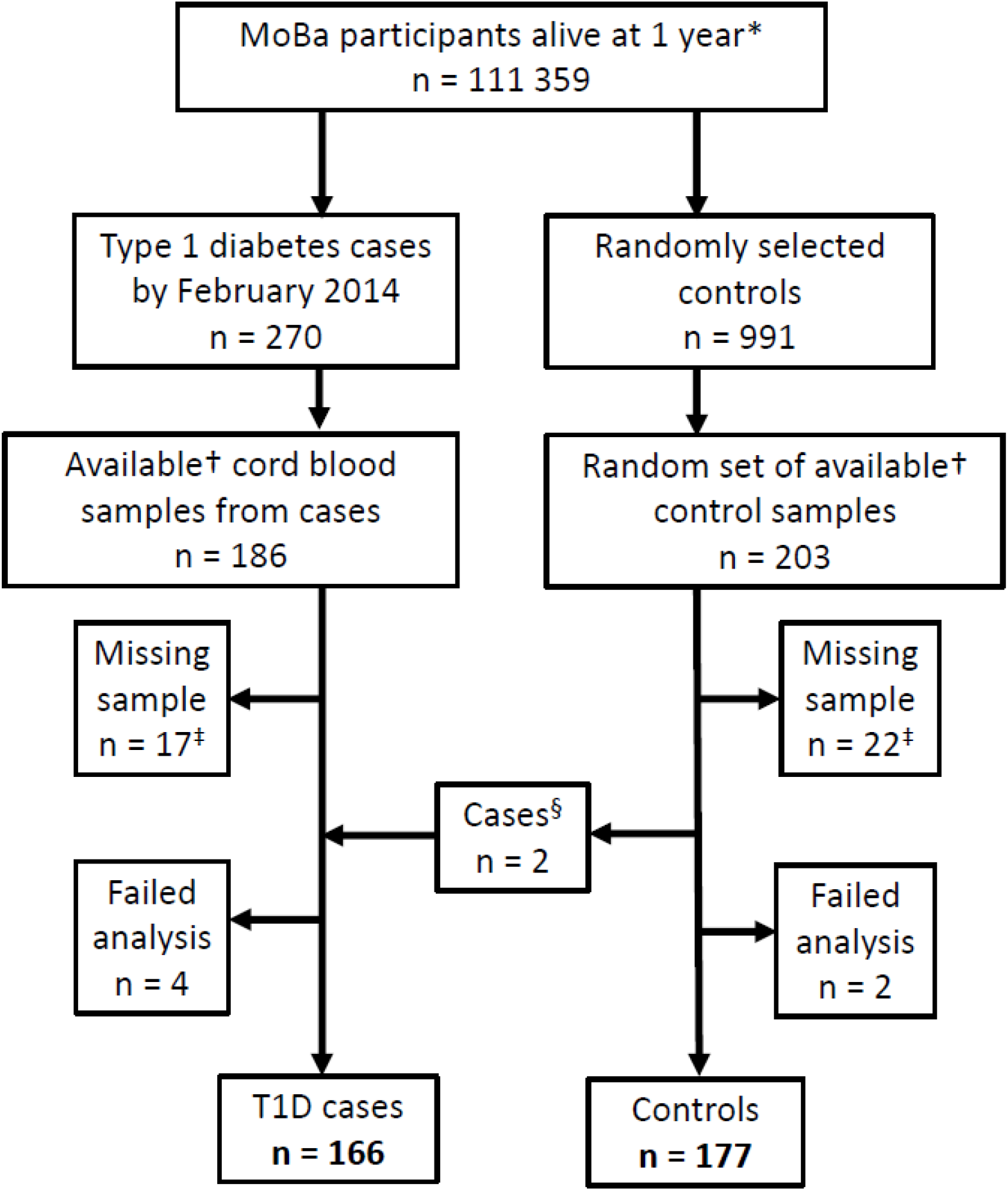
Flowchart. * Live-born children who survived their first year of life. ^†^ Cord blood samples registered, processed and not reserved for other projects. ^‡^ Includes cord blood samples with too low volume and unseparated blood. ^§^ We ascertained if any random control child developed type 1 diabetes by June 1, 2018 and these (n = 2) were reclassified as cases.

### Cord blood metabolomics profiling

Blood was sampled from the umbilical cord vein using a syringe immediately after birth, collected in a EDTA container and shipped to the biobank for plasma separation and storage at −80°C [30]. We used a UHPLC-QQQ-MS platform for targeted metabolomics with focus on robust measurement as described by Ahonen et al. [13]. Briefly, measurements were normalized, outliers were removed, then metabolites were excluded on the basis of high missingness (n=1) or low coefficient of variation (n=3) prior to analysis. Details are given in the Electronic Supplemental Material (ESM). In total, 27 metabolites were included for analysis, representing various metabolic pathways. A brief description of the selected metabolites is provided in ESM Table 1. The distributions of the plasma metabolite concentrations were approximately symmetric after log-transformation (ESM Figure 1). Z-scores were calculated to compare metabolites across different scales.

**Table 1:** Characteristics of participants.

|  | **Controls (n = 177)** | **Cases (n = 166)** |
| --- | --- | --- |
| **Age (median, range) at end of follow-up\* (years)** | 10.8 (7.1 – 15.7) | 12.0 (7.3 – 15.6)<sup>†</sup> |
| **Female sex** | 80 (45.2%) | 81 (48.8%) |
| **Preterm birth<sup>‡</sup>** | 5 (2.8%) | 10 (6.0%) |
| **Caesarean section<sup>§</sup>** | 12 (6.8%) | 28 (16.9%) |
| **Parity** |  |  |
| No earlier births | 77 (43.5%) | 83 (50.0%) |
| One | 63 (35.6%) | 49 (29.5%) |
| Two or more | 37 (20.9%) | 34 (20.5%) |
| **Maternal age (years (median, range))** | 30 (20 – 42) | 30 (19 – 42) |
| 19-24 | 24 (13.6%) | 14 (8.4%) |
| 25-34 | 128 (72.3%) | 130 (78.3%) |
| 35-42 | 25 (14.1%) | 22 (13.3%) |
| **Maternal smoking during pregnancy** |  |  |
| Non-smoker at end of pregnancy | 153 (86.4%) | 143 (86.1%) |
| Smoked at end of pregnancy | 17 (9.6%) | 18 (10.8%) |
| *Missing data* | 7 (4.0%) | 5 (3.0%) |
| **Pre-pregnancy BMI (kg/m<sup>2</sup> (median, range))** | 22.8 (17.2 – 40) | 23.9 (15.8 – 40.5) |
| <25 | 123 (69.5%) | 88 (53.0%) |
| 25-30 | 27 (15.3%) | 42 (25.3%) |
| >30 | 15 (8.5%) | 24 (14.5%) |
| *Missing data* | 12 (6.8%) | 12 (7.2%) |
| **Child's HLA genotype<sup>\*\*</sup>** |  |  |
| Protective (DQ6) | 55 (31.1%) | 4 (2.4%) |
| Neutral (any other HLA not mentioned) | 29 (16.4%) | 5 (3.0%) |
| Increased risk (≥1 copy of either DQ8 or DQ2.5) | 70 (39.5%) | 86 (51.8%) |
| High risk (DQ8/DQ2.5 heterozygote) | 11 (6.2%) | 67 (40.4%) |
| *Missing data* | 12 (6.8%) | 4 (2.4%) |

\* End of follow-up was set as the last diagnosis date of the cases, June 2, 2016
<sup>†</sup> Median age of diagnosis was 5.8 (range 0.7 – 12.8). only 3 cases were diagnosed after 12 years.
<sup>‡</sup> Defined as birth prior to pregnancy week 37.
<sup>§</sup> Includes elective (7 controls, 12 cases) and acute (5 controls, 16 cases) caesarean section.
<sup>||</sup> Including those that quit smoking shortly before or during pregnancy, as the protective association with type 1 diabetes has been observed in those that smoked throughout pregnancy. [48]
<sup>\*\*</sup> Coded as a binary variable in the analysis – protective or neutral (0) vs increased or high risk (1).
The increased risk category (≥1 copy of either DQ8 [DQA1\*03-DQB1\*03:02-DRB1\*04] or DQ2.5 [DQA1\*05:01-DQB1\*02:01] also required no HLA DQ6 [DQA1\*01:02-DQB1\*06:02-DRB1\*15:01] protective allele).

### Genotyping assays and genetic risk scores

Participants were genotyped for established type 1 diabetes susceptibilty SNPs including HLA tagSNPs using a custom Illumina Golden Gate assay (Illumina, San Diego, CA), as described earlier [31]. We calculated a 51 SNP non-HLA type 1 diabetes genetic risk score (GRS), weighted by the natural log-odds ratio of type 1 diabetes per risk allele reported from large GWAS studies (ESM Table 2, with further details given in the online supplement to [32]). Human leukocyte antigen (HLA) class II alleles were imputed using the HLA*IMP:02 web service [33] (details given in [31]) and all HLA genotypes were subsequently confirmed by allele specific PCR [34].

### Other covariates

Information on birth weight, maternal age at delivery, and delivery mode was obtained from the nationwide Medical Birth Registry of Norway [35]. Information regarding maternal pre-pregnancy body mass index (BMI) and smoking during pregnancy was obtained from mid-pregnancy and child’s 6 month of age questionnaires. Questionnaires can be accessed at www.fhi.no/moba. Maternal type 1 diabetes data were obtained from questionnaires and the Norwegian Patient Registry. Variables were categorized as shown in Table 1.

### Statistical methods

We used logistic regression with child type 1 diabetes as outcome, adjusted for selected variables, unless noted otherwise. The primary and secondary analyses were decided *a priori*. Our primary analysis was to estimate the logit-linear association between each single cord blood metabolite and child type 1 diabetes risk. Tertiles of each metabolite were used as categorical exposures to test our linearity assumption. We generated six clusters (groups of individuals with similar patterns across all metablites passing quality control) using k-means clustering, assessing whether the six clusters associated with type 1 diabetes risk, to investigate if any pattern of metabolites associated with type 1 diabetes.

Secondary analyses included restricting the primary analysis to children carrying type 1 diabetes risk HLA genotypes. We also evaluated the predictive performance of all the metabolites simultaneously, by calculating the area under the receiver operating characteristic curve (AUC) and compared with the AUC of well-established genetic type 1 diabetes susceptibility markers. A model including z-scores of all analysed metabolites, a model using HLA and a model using a weighted GRS were used to generate predictions for later type 1 diabetes based on metabolites, HLA, and non-HLA GRS, respectively. Five-fold crossvalidation was used in the AUC analysis to adjust for potential overfitting. We also investigated groups of metabolites by using the sum of amino acid levels, sum of bile acid levels, and the ratio of taurine/glycine-conjugated bile acids as exposures.

Exploratory analyses included principal component analysis (PCA) using principal components with an eigenvalue above 1 as predictors, and the MetaboAnalyst web service for metabolite set enrichment analysis (MSEA). [36] Details of the statistical analysis are given in ESM Methods.

Missing values were excluded in the analyses, with the exception of the cluster, AUC, PCA, enrichment, groups (amino and bile acids) and ratios of metabolites analyses, where we imputed the sample mean for missing values to allow inclusion of all metabolites. We used clustered sandwich estimator for standard errors to account for correlation between siblings. We present nominal p-values (not corrected for multiple testing), and 95% confidence intervals for the odds ratio excluding 1.00 were considered equivalent to a nominal significance at the 2-sided 5% level. We planned *a priori* to use false discovery rate and calculate q-values to account for multiple testing with 27 metabolites.

The following covariates were included in our primary adjustment model: sample batch, date of run, child’s sex, caesarean delivery, gestational age at birth (in weeks), maternal age, pre-pregnancy body mass index (BMI), smoking in pregnancy and parity (categorized as shown in Table 1). Analyses were run in R version 4.0.2, [37] and Stata release 16 (StataCorp LLC, College Station, TX).

## Results

Pairwise correlation coefficients among metabolites ranged from 0.77 for glycocholate-taurocholate and 0.62 for tyrosine-phenylalanine, to inverse correlations of −0.74 for glutamine-glutamic acid and −0.55 for taurine-glutamine, but most metabolite pairs showed little correlation (ESM Figure 2).

**Figure 2:**
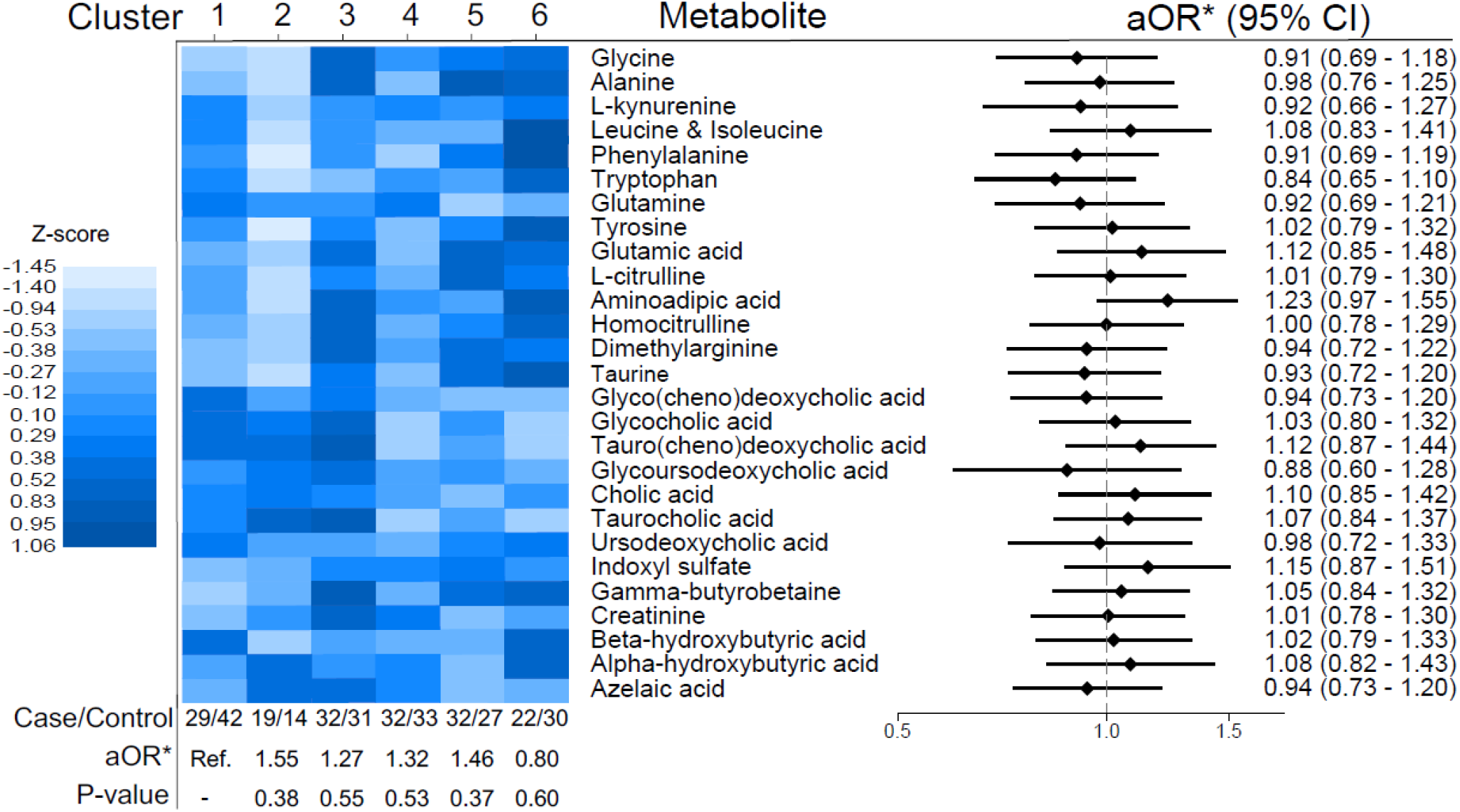
Results from cluster analysis and logistic regression. aOR: adjusted Odds Ratio Figure 2 shows the results from the cluster analysis (with a heatmap representing mean metabolite concentrations in each cluster) and the results from the main analysis (logistic regression, with each metabolite in a separate model) as a forest plot. *Adjusted for sample batch, date of run, child’s sex, caesarean delivery, length of pregnancy (in weeks), maternal age, pre-pregnancy body mass index (BMI), smoking in pregnancy and parity.

### Main analysis: Association of individual metabolites and clusters with type 1 diabetes

The strongest associations were observed for aminoadipic acid (aOR=1.23, 95%CI:0.97–1.55), indoxyl sulfate (aOR=1.15, 95%CI:0.87–1.51), taurochenodeoxycholic acid (aOR=1.12, 95%CI:0.87–1.44), and tryptophan (aOR=0.84, 95%CI:0.65–1.10), with other aORs close to 1.0. No metabolite was nominally significantly associated with type 1 diabetes (Figure 2). Categorizing metabolites into tertiles did not show deviation from linearity, except for phenylalanine (ESM Figure 3). The second tertile of phenylalanine was associated with lower risk of type 1 diabetes, while the lowest and highest tertile had neutral risk (ESM Figure 3), meaning the linear estimate reported must be cautiously interpreted. Adjusting for batch, date of run and maternal BMI or birthweight only, variables which could be the most relevant for metabolite levels and offspring type 1 diabetes, did not appreciably change our results (data not shown). When individuals were clustered using k-means clustering into six groups of cord blood metabolite profiles, no cluster was significantly associated with type 1 diabetes (Figure 2).

**Figure 3.**
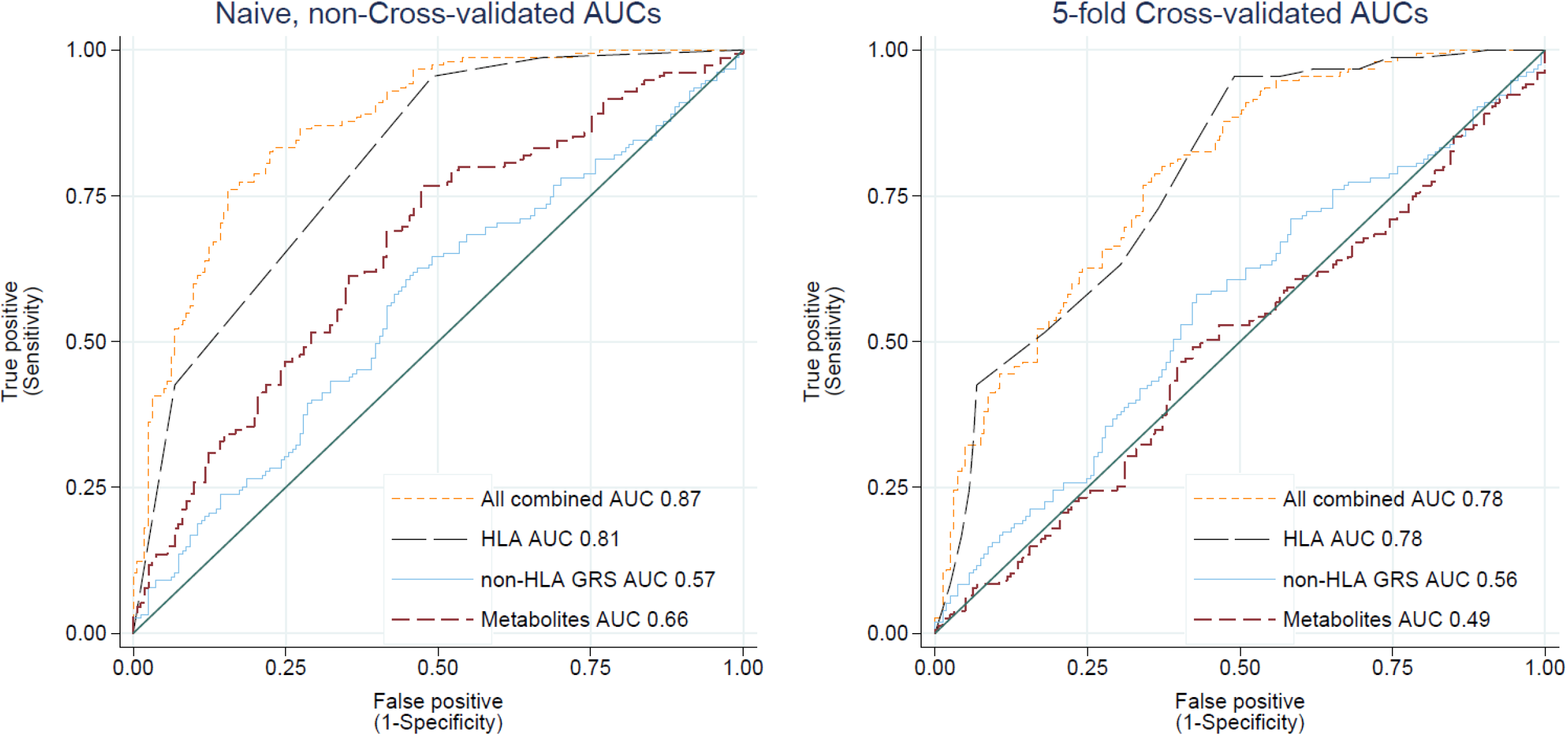
Area under the receiver operating characteristics curves (AUC) of predicted risk of type 1 diabetes, using predictions from logistic regression with either 27 cord blood metabolites, a 51 SNP non-HLA genetic risk score (GRS), 4-category HLA risk group, or all combined, both cross-validated (CV) and not.

### Secondary analyses: Multivariable prediction of type 1 diabetes at birth with ROC curves

Using all 27 metabolites to predict later type 1 diabetes with multiple logistic regression gave an AUC of 0.66, the non-HLA genetic risk score alone an AUC of 0.57, and HLA risk group alone gave an AUC of 0.81, and all combined an AUC of 0.87 before cross validation (Figure 3a). Because the non-HLA SNP was externally weighted, it was not expected to give a too optimistic prediction (overfitting), whereas the HLA and metabolite predictions were not externally weighted. After accounting for potential overfitting using five-fold cross-validation, the metabolites showed no predictive value, while the HLA group and non-HLA genetic risk score were only marginally attenuated after cross validation. HLA remained the clearly most important predictor (Fig 3b). This illustrates that the initial suggestive predictive utility of metabolites was likely due to overfitting.

### Other secondary analyses

Analysing metabolite groups (by summing the z-scores of amino acids, or bile acids, and using these variables as exposures) did not show any statistical association with later type 1 diabetes (amino acids aOR=1.00, 95%CI:0.96–1.03, bile acids aOR=1.02, 95%CI:0.95–1.09). The taurine/glycine-conjugated bile acid ratio was not associated with offspring type 1 diabetes (aOR=1.07, 95%CI:0.91–1.26). Restricting our analysis to only children carrying HLA genotypes conferring increased type 1 diabetes risk gave similar results as the main analysis (ESM Table 3).

### Exploratory analyses

Metabolite Set Enrichment Analysis did not show any statistically significant results (ESM Figure 4). PCA analysis of the metabolites detected eight principal components with eigenvalues larger than 1. The cumulative proportion of variance of the components was 0.62, and none of these components were associated with later type 1 diabetes (ESM Figure 5). Pairwise plotting of the principal components did not show any clear pattern (an example is shown in ESM Figure 6).

## Discussion

In this study, we investigated selected small polar metabolites measured in cord blood and their potential association with later type 1 diabetes. No metabolites, or cluster of individuals with different metabolite profile, were significantly associated with later type 1 diabetes.

### Comparison to other studies

To our knowledge, there are no previous studies on maternal or infant bile acids and their potential association with type 1 diabetes. There are only two studies on polar metabolites at birth and type 1 diabetes. Orešič et al. reported lower levels of several metabolites, including tryptophan, glycine and creatinine, in cord blood associated with later type 1 diabetes in the Finnish DIPP cohort. [24] La Marca et al. reported lower levels of alanine and carnitines in children who later developed type 1 diabetes compared to children who did not, using dried blood spots taken shortly after birth. [25] Our findings are not consistent with results from these studies, but we did not measure carnitines. Cord blood and dried blood spots might also not be directly comparable.

Metabolomics during infancy and early childhood and association with islet autoimmunity or type 1 diabetes have been investigated in a few previous studies The DIPP cohort reported lower glutamic acid and tryptophan at 3 or 6 months of age (q-values <0.1), [38] and lower concentrations of branched chained amino acids, phenylalanine and tyrosine in peripheral blood mononuclear cells (PBMCs) at 12 months of age (not significant after multiple testing correction), in children who developed type 1 diabetes compared to controls. [39] In the international TEDDY cohort, Johnson et al., measured metabolites at 9 months of age, and reported dicarboxylic acids (adipic acid being most significant) to be associated with increased risk of subsequent islet autoimmunity. [40] Li et al., also based on the TEDDY cohort, reported alanine and β-hydroxybutyric acid, at 12 and 24 months of age, respectively, to be associated with lower risk of developing islet autoimmunity in those developing autoantibodies against insulin first. [41] The Norwegian MIDIA study reported differences in amino acid levels, albeit not statistically significant after correction for multiple testing, [42] and the German BABYDIAB study reported lower methionine levels, [43] after development of islet autoantibodies. There are also two publications from TEDDY and one from DAISY using polar metabolites to predict later islet autoimmunity. Stanfill et al. used classification algorithms to determine the most predictive features of islet autoimmunity in the TEDDY study (504 samples) and reported adipic acid, creatinine, and leucine as influential metabolites. [44] Webb-Robertson et al. used a machine learning approach to predict islet autoimmunity using data from the TEDDY study (157 case-control pairs), reporting azelaic acid and adipic acid as important features. [45] Frohnert et al. predicted seroconversion to islet autoimmunity in the DAISY study (22 cases and 25 controls) and reported 3-methyl-oxobutyrate (a precursor to valine for leucine synthesis) and pyroglutamatic acid (a derivative of glutamic acid) as features that were often selected by the algorithm used. [46]

We did not find any strong associations between specific metabolites measured at birth and later type 1 diabetes. Likewise, we did not find any cluster or group of metabolites that were overrepresented in children who later developed type 1 diabetes, and the metabolites studied had low predictive value for later type 1 diabetes. It must be kept in mind that earlier studies are heterogenous in most aspects: sample handling, measurement method, exposures measured, results reported, statistical analysis and endpoint (for an overview, see ESM Table 4), which makes results difficult to compare. Few studies adjusted for covariates but adjusting for covariates or stratifying by type 1 diabetes HLA risk did not change our estimates to a large degree. Approaches to control multiplicity varies and it is not always clear how the problem has been handled in previous studies. It is not known if participants/samples included in subsequent publications from the same cohort (such as publications from the DIPP and TEDDY cohorts) overlap to a large degree or are separate. With the exception of Orešič et al. [24], and la Marca et al., [25] other studies have not measured polar metabolites at birth. There could also be differences between countries, or the population studied that lead to different results.

The previously published prediction studies using postnatal circulating metabolites are likewise not directly comparable, and there are few consistent findings. The reported AUCs differ across studies, as does the data analysis and validation approach. Frohnert et al., starting with 1552 features (and 22 cases), reported an AUC of 0.92 for prediction of islet autoimmunity in the DAISY study, without a holdout validation set. [46] Webb-Robertson et al., using the TEDDY cohort, reported that the top 42 features (of 221) gave an AUC of 0.65 for prediction of islet autoimmunity in the holdout validation set, appreciably lower than the AUC of 0.74 estimated with cross-validation of the training set. [45] As seen in our results, cross-validation is essential to avoid overfitting, even when using few features and a relatively large sample size (166 cases, 177 controls). Several factors influence the potential for overfitting and bias, but in general, the larger the number of predictors in relation to the number of subjects with outcome, and the more flexible modelling approach (allowing non-linearities and interactions), the larger the potential for overfitting. Using a non-validated AUC analysis could lead to the misinterpretation that these metabolites have predictive value for later type 1 diabetes in the same range as well-established genetic risk factors, and would add to the predictive value of genetic factors. Cross-validation shows that this overly optimistic results is due to overfitting even with a simple logistic regression model, while well-established genetic factors had an essentially unchanged AUC. This is consistent with the fact that overfitting bias tend to be smaller the greater the *a priori* evidence for the predictors used.

While our approach was to start with a limited number of predictors (n=27) in the model without statistical selection, an approach using statistical selection, such as stepwise selection or least absolute shrinkage and selection operator (LASSO), would not select any metabolites and thus not obtain an overfitted model. We nevertheless think our results provide an important message regarding potential bias in multivariable prediction of type 1 diabetes or other outcomes. While overfitting and multiple testing are related concepts, reducing the influence of these requires different techniques. Overfitting is a problem of multivariable prediction and can be minimised by cross validation and independent validation data. For correcting multiple statistical testing, we had planned to control the false discovery rate at 5%, and compute q-values. As no single metabolite was significant at the nominal 5% level, we conclude that no metabolite was significant, and q-values do not make any sense in this case. [47]

### Interpretation and implication of results

Our goal was to investigate if metabolites at birth could be predictive of later type 1 diabetes, by studying selected metabolites both singly and combined, and investigating their predictive value. Our interpretation of the data is that these metabolites, measured in cord blood, are of limited importance in offspring type 1 diabetes. The evidence that intrauterine exposures are associated with type 1 diabetes is not extensive. [3] This does not mean that no metabolic changes occur closer to development of disease, but these possible differences do not seem present at birth for the measured metabolites. Lack of predictive value of these metabolites also does not negate the existence of other perinatal exposures that may influence the risk of type 1 diabetes, if we assume such factors are not strongly correlated with the metabolites measured in our study. The relatively consistently replicated associations between maternal obesity, birth weight and type 1 diabetes could exert their potential effects through other means.

### Strengths and limitations

A strength of the study is the use of cord blood samples, which allows us to study exposures before onset of autoimmunity and dysglycaemia, and characterize influences during intrauterine life, including potential maternal exposures in pregnancy. We include children from the general population, not selected by family history or HLA risk genotypes, which makes our results more applicable to the general population. This study is the largest measuring polar metabolites in cord blood. The targeted approach allowed us to quantify a set of metabolites with a biological basis. This reduce problems of multiplicity (multiple testing and overfitting), with the potential cost of not including potentially relevant metabolites. Still, increasing the number of measured metabolites is likely to offset potential gains by increasing multiplicity problems. As with any observational study, we cannot rule out unmeasured confounders, although we do not believe unmeasured confounders have changed our conclusions.

## Conclusions

In this large study, the selected polar metabolites measured in cord blood were not associated with later type 1 diabetes in the offspring.

## Supporting information

Electronic Supplementary Material

## Data Availability

Data availability: Aggregated data is available from the authors upon reasonable request. The consent given by the participants does not open for storage of data on an individual level in repositories or journals. Access to individual level data sets requires an application, approval from The Regional Committee for Medical and Health Research Ethics in Norway and an agreement with MoBa.

## Acknowledgements

The Norwegian Mother, Father and Child Cohort Study is supported by the Norwegian Ministry of Health and Care Services and the Ministry of Education and Research. We are grateful to all the participating families in Norway who take part in this on-going cohort study. The Norwegian Childhood Diabetes Registry is funded by The South-Eastern Norway Regional Health Authority. We are thankful to the Norwegian Childhood Diabetes Registry and Study Group. Costs of all data acquisition, including laboratory assays in MoBa (the sub-study PAGE; Prediction of Autoimmune Diabetes and Celiac Disease in Childhood by Genes and Perinatal Environment), was supported by grant 2210909/F20 from the Norwegian Research Council (Dr. Lars C. Stene). Supported by grants (to Dr. Njølstad) from the European Research Council (AdG #293574), the Bergen Research Foundation (“Utilizing the Mother and Child Cohort and the Medical Birth Registry for Better Health”), Stiftelsen Kristian Gerhard Jebsen (Translational Medical Center), the University of Bergen, the Research Council of Norway (FRIPRO grant #240413), the Western Norway Regional Health Authority (Strategic Fund “Personalized Medicine for Children and Adults”), the Novo Nordisk Foundation (grant #54741), and the Norwegian Diabetes Association. Data from the Norwegian Patient Register has been used in this publication. The interpretation and reporting of these data are the sole responsibility of the authors, and no endorsement by the Norwegian Patient Register is intended nor should be inferred. Genotyping in this study was performed at the Genomics Core Facility, Norwegian Radium hospital, Oslo, Norway. We are thankful to Dr. K. Gillespie and Miss G. Mortimer (Diabetes and Metabolism, School of Clinical Sciences, Southmead Hospital, University of Bristol, Bristol, UK) for the HLA typing. In addition, the authors thank laboratory technician Birgitte Nergaard Roberts (Steno Diabetes Center Copenhagen), who was responsible for the extensive sample randomization and sample preparation before the UHPLC-QQQ-MS analyses.

## Data availability

Aggregated data is available from the authors upon reasonable request. The consent given by the participants does not open for storage of data on an individual level in repositories or journals. Access to individual level data sets requires an application, approval from The Regional Committee for Medical and Health Research Ethics in Norway and an agreement with MoBa.

## Author contributions

Conception and design: GT, LCS, KS.

Literature search: GT, LCS

Acquisition of pregnancy cohort data: LCS, KS.

Acquisition of incident type 1 diabetes data: TSk, GJ, PRN.

Measurement of cord blood metabolites: TSu, LA, CLQ.

Data cleaning and preparation: GT, TSu.

Planning statistical analyzes: LCS, GT, KS.

Performing statistical analyses: GT.

Interpretation of data: All authors.

Drafting the manuscript: GT.

Revising the manuscript critically for important intellectual content: All authors.

Final approval of the version to be published: All authors

Taking responsibility for the integrity of the data and the accuracy of the data analysis: GT, LCS.

Obtaining funding: LCS, KS.

