## Supplementary material for "Prediction of type 1 diabetes at birth: cord blood metabolites versus genetic risk score in the MoBa cohort": Electronic Supplementary Material

Affiliations:
[1] Department of Chronic Diseases and Ageing, Norwegian Institute of Public Health, Oslo, Norway
[2] Steno Diabetes Center, Copenhagen, Denmark
[3] Biosyntia ApS, Copenhagen, Denmark
[4] Division of Pediatric and Adolescent Medicine, Oslo University Hospital, Oslo, Norway
[5] Department of Pediatric and Adolescent Medicine, Haukeland University Hospital, Bergen, Norway
[6] Center for Diabetes Research, Department of Clinical Science, University of Bergen, Bergen, Norway
[7] Institute of Clinical Medicine, University of Oslo, Oslo, Norway
[8] Pediatric Research Institute, Institute of Clinical Medicine University of Oslo, Oslo, Norway

Corresponding author: German Tapia, Department of Chronic Diseases and Ageing, Norwegian Institute of Public Health, P.O. Box 222 Skøyen, NO-0213 Oslo, Norway.

### Electronic Supplementary Material (ESM) Methods

We used a targeted clinical metabolite profiling platform, as described by Ahonen et al. which measures 34 selected metabolite biomarkers from plasma. [1] Few pairs of metabolites with high chemical similarity had similar molecular mass and fragmentation pattern, resulting in overlapping peaks and were not chromatographically separable. These metabolite pairs were quantified together(L-Leucine and L-Isoleucine, Taurodeoxycholic and Taurochenodeoxycholic acid, Glycodeoxycholic and Glycochenodeoxycholic Acid, Asymmetric and Symmetric dimethylarginine, Table 1). [1] For some metabolites, quantification failed, which could be due to low levels in the samples, or the calibration not meeting our required accuracy. For these metabolites (L-Homocitrulline, Glycoursodeoxycholic acid, Hydroxybutyric acid, Indoxyl sulfate, Creatinine and Azelaic acid), normalized peaks were used for the statistical analysis. For few metabolites (Glutamine, Glycocholic acid, Taurodeoxycholic and Taurochenodeoxycholic acid), technical variation did not reduce upon normalization to the respective internal standard. Therefore, original peak areas were used to calculate exposure In the statistical analysis.

First, clearly implausible values (at least 20 times higher than the next-highest value; n=4) were set to missing. Metabolites were log2 transformed, followed by excluding outliers, defined as observations being six times above or below the median absolute deviation (MAD; defined as the median of the absolute deviations from the variable median). [2] Metabolites with more than 20% missing (Deoxychenocholic acid), and metabolites with a relative standard deviation <5% (the standard deviation divided by the mean, also known as the coefficient of variation: Deoxycholic acid, N-methylnicotinamide and Tauroursodeoxycholic acid) were exluded.

*Metabolite set enrichment analysis.*

We used metabolite set enrichment analysis (MSEA) to investigate if any metabolic pathway was perturbed at birth in children who later developed type 1 diabetes. MSEA uses predefined metabolite sets covering various metabolic pathways and analyses the metabolites as groups (e.g. cysteine metabolism or fatty acid synthesis), giving the mean fold difference of the metabolic pathways in children who later developed type 1 diabetes. Metabolites are grouped on the basis of being involved in the same biological processes. [3] We performed a quantitative enrichment analysis, which is based on the *globaltest* algorithm,[4] and uses a linear model to estimate a "Q-stat" for each metabolite set. [3] The "Q-stat" describes the correlation between the metabolite concentrations in a set and outcome. [3] MSEA is available online with the MetaboAnalyst web-based metabolomics tool suite. [5] For details see Xia et al. [3] and Chong et al. [5].

### ESM Table 1: Overview of metabolites studied

| Metabolite | Abbr. | Quantification status | Biological background |
|---|---|---|---|
| ***Amino Acids and related metabolites*** | | | Several studies report amino acid differences could possibly be associated to type 1 diabetes or IA. The placenta actively transports most amino acids. Most amino acids and their metabolites (except asparagine/aspartate, and arginine) showed significant correlation between maternal fasting plasma around gestational week 28, and cord blood levels. [6] |
| Glycine | Gly | Concentration | Lower serum glycine associated with risk of impaired glucose tolerance and type 2 diabetes. [7] Reported lower at birth in those that later develop islet autoimmunity by Orešič et al. [8] |
| L-Alanine | Ala | Concentration | Increased levels of alanine linked to development of type 2 diabetes. [9, 10] Lower levels reported at birth in children who later develop type 1 diabetes. [11] |
| L-Leucine and L-Isoleucine | Leu/Ile | Concentration | Associated with insulin resistance and type 2 diabetes. [10, 12] |
| L-Phenylalanine | Phe | Concentration | Stimulates insulin release. Linked to insulin resistance change in amino acid profile, and increased T2D risk (several studies, reviewed in [12]). |
| L-Tryptophan | Trp | Concentration | Tryptophan metabolism considered important for regulating T cell immunity. [13] Lower levels possibly associated with development of islet autoimmunity. [8, 14, 15] |
| L-Glutamine | Gln | Peak area | Associated to lower risk of type 2 diabetes. [12] Glutamine is involved in multiple processes that may impact insulin sensitivity and/or glucose homeostasis.[16] |
| L-Tyrosine | Tyr | Concentration | Lower levels at 12 months of age in children who later developed type 1 diabetes. [17] Linked to increased type 2 diabetes risk (several studies, reviewed in [12]). |
| L-Glutamic Acid | Glu | Concentration | Possible association with islet autoimmunity in early childhood. [14, 15] |
| L-Citrulline | Cit | Concentration | Possible marker of acute and chronic small bowel cell mass.[18] Associated with prediabetes and type 2 diabetes. [12] |
| L-Homocitrulline | Hcit | Normalized peak area | Generated from Lysine reacting with cyanate. Possible marker of inflammation, as is generated during inflammation. Metabolite of ornithine. |
| DL-2-Aminoadipic acid | AADA | Concentration | Dicarboxylic acid associated with insulin resistance and beta cell function.[12, 19] |
| L-kynurenine | Kynu | Concentration | Higher in type 1 diabetes patients after diagnosis. [20] Synthesized as a result of immune activation, metabolite of tryptophan. [21] |
| Asymmetric & Symmetric dimethylarginine | A(S)DMA | Concentration | Levels inversely correlated with HbA(1c) concentrations and serum glucose levels. [22, 23] Markers of renal function. [24] |
| Taurine | Taur | Concentration | Abundant metabolite, conjugated with bile acids. Protective of diabetes in NOD mice. [25] Associated with lower type 2 diabetes risk. [26] |
| ***Bile Acids*** | | | Bile acids are important endocrine signaling molecules, modulating glucose homeostasis by regulating lipid, glucose and energy metabolism. [27, 28] Bile acids associated with insulin resistance. [29] Bile acids are transported from the fetus to the mother. [30] Fiber intake inversely correlated to bile acids. [31] Bile acids involved in gastrointestinal immunity. [32] |

| | | | |
|---|---|---|---|
| Glycochenodeoxycholic & Glycodeoxycholic acid | G(C)DCA | Concentration | Bile salt formed in the liver from deoxycholic/ chenodeoxycholic acid and glycine, involved in fat absorption. |
| Glycocholic acid | GCA | Peak area | Conjugate of cholic acid with glycine. |
| Taurochenodeoxycholic & Taurodeoxycholic acid | T(C)DCA | Peak area | Conjugate of chenodeoxycholic/deoxycholic acid with taurine. ER stress inhibitor. [33] |
| Glycoursodeoxycholic acid | GUDCA | Normalized peak area | Conjugate of ursodeoxycholic acid with glycine. |
| Cholic acid | CA | Concentration | Primary bile acid secreted by the liver, metabolized by bacteria to deoxycholic acid. |
| Taurocholic acid | TCA | Concentration | Conjugate of cholic acid with taurine. |
| Ursodeoxycholic acid | UDCA | Concentration | Secondary bile acid produced in humans by intestinal bacteria. Ursodeoxycholic acid reduce colitogenic dysbiosis in mice. [34] |
| ***Other metabolites*** | | | |
| Indoxyl sulfate | IndS | Peak area | Gut bacteria metabolite of tryptophan. Marker of kidney dysfunction. |
| γ-Butyrobetaine | GBB | Concentration | γ-Butyrobetaine is hydroxylated to produce carnitine, which has been reported lower in children who later developed type 1 diabetes at birth. [11] |
| Creatinine | Crea | Normalized peak area | Associated with lower type 2 diabetes risk. [10] Possible association with islet autoimmunity. [8, 35] Creatinine measured in cord blood largely reflects maternal levels. [36] |
| ***Small organic acids*** | | | |
| L-3-hydroxybutyric acid (β) | β-OHB | Concentration | Ketone body, increased during ketosis. Linked to early metabolic changes in T2D. [37] |
| R/S-2-hydroxybutyric acid (α) | α-OHB | Normalized peak area | Oxidative stress or detoxification can increase levels. Linked to early metabolic changes in T2D.[37] Linked to insulin resistance and impaired glucose regulation. [38] |
| Azelaic acid | AzelA | Normalized peak area | Dicarboxylic acid found naturally in wheat, rye and barley. Possible anti-obesity effects, reducing adiposity. Increased insulin sensitivity. Shifting to use fats for energy. Increase lipolysis and fatty acid oxidation. [39, 40] |

ESM Table 2: Non-HLA type 1 diabetes risk SNPs used in the GRS

| SNP | Gene(s) or position | Ln(OR) | OR | CHR | Ref. |
|---|---|---|---|---|---|
| rs2476601 | PTPN22 | 0.64 | 1.89 | 1 | [41] |
| rs2816316 | RGS1 | 0.12 | 1.12 | 1 | [42] |
| rs3024505 | IL10 | 0.17 | 1.19 | 1 | [42] |
| rs2111485 | IFIH1 | 0.16 | 1.18 | 2 | [41] |
| rs3087243 | CTLA4 | 0.17 | 1.19 | 2 | [41] |
| rs478222 | EFR3B | 0.14 | 1.15 | 2 | [43] |
| rs6740838 | AFF3 | 0.11 | 1.12 | 2 | [43] |
| rs7574865 | STAT4 | 0.06 | 1.06 | 2 | [43] |
| rs917997 | IL18RAP, IL18R1, IL1RL1, IL1RL2 | 0.02 | 1.02 | 2 | [44] |
| rs11711054 | CCR5 | 0.06 | 1.06 | 3 | [43] |
| rs10517086 | Position: 26083889 | 0.09 | 1.09 | 4 | [42] |
| rs4505848 | KIAA1109, IL2 | 0.11 | 1.12 | 4 | [43] |
| rs11755527 | BACH2 | 0.12 | 1.13 | 6 | [42] |
| rs1738074 | TAGAP | 0.08 | 1.09 | 6 | [42] |
| rs6920220 | TNFAIP3 | 0.11 | 1.12 | 6 | [41] |
| rs924043 | Position: 170063801 | 0.17 | 1.19 | 6 | [43] |
| rs9388489 | CENPW | 0.16 | 1.17 | 6 | [42] |
| rs10272724 | IKZF1 | 0.14 | 1.15 | 7 | [45] |
| rs4948088 | COBL | 0.26 | 1.30 | 7 | [42] |
| rs7804356 | SKAP2 | 0.13 | 1.14 | 7 | [42] |
| rs7020673 | GLIS3 | 0.13 | 1.14 | 9 | [42] |
| rs10509540 | C10orf59 | 0.29 | 1.33 | 10 | [42] |
| rs61839660 | IL2RA | 0.48 | 1.61 | 10 | [42] |
| rs947474 | PRKCQ | 0.10 | 1.10 | 10 | [46] |
| rs689 | INS | 0.87 | 2.38 | 11 | [41] |
| rs10492166 | CLECL1 | 0.14 | 1.15 | 12 | [43] |
| rs1265564 | CUX2 | 0.37 | 1.45 | 12 | [47] |
| rs17696736 | NAA25 | 0.29 | 1.34 | 12 | [48] |
| rs2292239 | ERBB3 | 0.25 | 1.28 | 12 | [49] |
| rs3764021 | CLEC2D | 0.45 | 1.57 | 12 | [48] |
| rs653178 | ATXN2 | 0.26 | 1.30 | 12 | [41] |
| rs917911 | CD69 | 0.10 | 1.10 | 12 | [41] |
| rs9585056 | EBI2 (GPR183) | 0.14 | 1.15 | 13 | [50] |
| rs1465788 | ZFP36L1 | 0.15 | 1.16 | 14 | [42] |
| rs4900384 | Position: 98032614 | 0.09 | 1.09 | 14 | [42] |
| rs941576 | DLK1 MEG3 | 0.11 | 1.11 | 14 | [51] |
| rs12908309 | RASGRP1 | 0.16 | 1.18 | 15 | [43] |
| rs3825932 | CTSH | 0.15 | 1.16 | 15 | [42] |
| rs12927355 | CLEC16A | 0.20 | 1.22 | 16 | [41] |
| rs4788084 | IL27 | 0.15 | 1.16 | 16 | [42] |
| rs8056814 | CTRB1 | 0.28 | 1.32 | 16 | [41] |
| rs2290400 | GSDMB | 0.14 | 1.15 | 17 | [42] |
| rs7221109 | SMARCE1 | 0.05 | 1.05 | 17 | [42] |
| rs1893217 | PTPN2 | 0.19 | 1.21 | 18 | [41] |
| rs763361 | CD226 | 0.15 | 1.16 | 18 | [42] |
| rs601338 | FUT2 | 0.25* | 1.14* | 19 | [52] |
| rs2281808 | SIRPG | 0.11 | 1.11 | 20 | [42] |
| rs11203203 | UBASH3A | 0.13 | 1.14 | 21 | [43] |
| rs229541 | C1QTNF6 | 0.10 | 1.11 | 22 | [46] |
| rs2412970 | HORMAD2 | 0.09 | 1.10 | 22 | [43] |
| rs2664170 | GAB3 | 0.15 | 1.16 | 23 | [42] |

* Calculated per increase in risk allele from estimates given in Smyth et al [52].

SNP: single nucleotide polymorphism
CHR: chromosome

ESM Table 2: This table shows the non-HLA SNPs, genes near these (or position) and the weights (the natural log of the odds ratios [ORs]) used to calculate the weighted genetic risk score (GRS) used in our study. The GRS was calculated by multiplying the number of risk alleles with the natural logarithm of the OR per each SNP, then summing this for each individual. More details are provided in the supplement to Vistnes et al.[53]

### ESM Table 3: Sensitivity analysis

| Metabolite | Not adjusted for covariates | | HLA-restricted analysis | |
|---|---|---|---|---|
| | aOR*, 95%CI | p | aOR†, 95%CI | p |
| Glycine | 1.00 (0.79 - 1.27) | 0.98 | 1.01 (0.75 - 1.35) | 0.95 |
| Alanine | 1.06 (0.85 - 1.32) | 0.60 | 1.02 (0.79 - 1.32) | 0.90 |
| L-kynurenine | 1.03 (0.78 - 1.35) | 0.84 | 1.02 (0.70 - 1.46) | 0.93 |
| Leucine & Isoleucine | 1.01 (0.80 - 1.27) | 0.92 | 0.99 (0.74 - 1.33) | 0.97 |
| Phenylalanine | 0.95 (0.75 - 1.21) | 0.68 | 0.84 (0.61 - 1.16) | 0.28 |
| Tryptophan | 0.81 (0.63 - 1.04) | 0.10 | 0.83 (0.60 - 1.14) | 0.25 |
| Glutamine | 0.89 (0.70 - 1.13) | 0.33 | 0.83 (0.60 - 1.16) | 0.28 |
| Tyrosine | 1.06 (0.84 - 1.34) | 0.64 | 0.87 (0.63 - 1.21) | 0.41 |
| Glutamic acid | 1.11 (0.87 - 1.41) | 0.40 | 1.13 (0.82 - 1.54) | 0.46 |
| L-citrulline | 1.06 (0.85 - 1.33) | 0.61 | 1.01 (0.76 - 1.35) | 0.94 |
| Aminoadipic acid | 1.19 (0.96 - 1.48) | 0.10 | 1.27 (0.97 - 1.67) | 0.09 |
| Homocitrulline | 1.09 (0.87 - 1.37) | 0.47 | 1.05 (0.78 - 1.42) | 0.75 |
| Dimethylarginine | 1.06 (0.83 - 1.34) | 0.65 | 0.98 (0.71 - 1.35) | 0.91 |
| Taurine | 0.91 (0.72 - 1.15) | 0.44 | 0.90 (0.66 - 1.22) | 0.50 |
| Glyco(cheno)deoxycholic acid | 0.85 (0.68 - 1.07) | 0.17 | 0.79 (0.60 - 1.03) | 0.08 |
| Glycocholic acid | 1.00 (0.79 - 1.26) | 0.99 | 0.94 (0.71 - 1.26) | 0.69 |
| Tauro(cheno)deoxycholic acid | 1.14 (0.92 - 1.43) | 0.23 | 1.01 (0.76 - 1.34) | 0.95 |
| Glycoursodeoxycholic acid | 0.97 (0.69 - 1.36) | 0.86 | 1.15 (0.76 - 1.75) | 0.50 |
| Cholic acid | 1.03 (0.82 - 1.29) | 0.81 | 1.03 (0.76 - 1.41) | 0.85 |
| Taurocholic acid | 1.06 (0.84 - 1.33) | 0.63 | 1.01 (0.76 - 1.34) | 0.94 |
| Ursodeoxycholic acid | 1.12 (0.86 - 1.47) | 0.41 | 1.26 (0.88 - 1.79) | 0.20 |
| Indoxyl sulfate | 1.06 (0.83 - 1.36) | 0.66 | 1.03 (0.76 - 1.41) | 0.84 |
| γ-butyrobetaine | 1.13 (0.93 - 1.39) | 0.23 | 1.04 (0.82 - 1.32) | 0.75 |
| Creatinine | 1.02 (0.82 - 1.28) | 0.85 | 0.98 (0.73 - 1.32) | 0.91 |
| β-hydroxybutyric acid | 0.94 (0.75 - 1.18) | 0.62 | 0.96 (0.72 - 1.26) | 0.76 |
| α-hydroxybutyric acid | 1.09 (0.85 - 1.40) | 0.48 | 1.15 (0.85 - 1.55) | 0.37 |
| Azelaic acid | 0.95 (0.76 - 1.18) | 0.64 | 0.86 (0.64 - 1.16) | 0.32 |

OR: Odds Ratio; aOR: adjusted Odds Ratio

* Adjusted only for sample batch and date of run.

† Adjusted only for sample batch and date of run, including only participants with a HLA genotype conferring increased type 1 diabetes risk (≥1 copy of either DQ8 [DQA1*03-DQB1*03:02-DRB1*04] or DQ2.5 [DQA1*05:01-DQB1*02:01], and no protective HLA DQ6 [DQA1*01:02-DQB1*06:02-DRB1*15:01] allele) in the analysis. This analysis includes 153 cases and 81 controls.

ESM Table 3 shows the results from two sensitivity analyses; ananalysis adjusted only for sample batch and preparation date, and an analysis only including participant with a genotype conferring HLA risk.

#### ESM Table 4: Overview of earlier studies of plasma or serum metabolomics in type 1 diabetes

| Author, year | Cohort | Cases | Controls | Matching | Adjustment | Summary of most relevant results | Earliest measurement at |
|---|---|---|---|---|---|---|---|
| la Marca, 2013 [11] | Hospital-based* | 50 type 1 diabetes | 200 | No matching, random controls born +/- 1 day of case | No | Lower carnitines and alanine in children who later developed type 1 diabetes. “Trend” of lower amino acids in children who later developed type 1 diabetes. | Routinely collected dried blood spots, taken 48-72 hours after birth (analyzed within 2 days of collection). |
| Orešič, 2008 [8] | DIPP | 15 type 1 diabetes | 24 | 1:1 Birth season, Birth city, Sex, HLA | No | Lower α-ketogluaric acid, Tryptophan, Serine, Glycine, Ribitol, Succinic acid, Phosphoric acid, Lactic acid, Ornithine, Creatinine and Citric acid | Birth (cord blood serum) |
| Lamichhane, 2019 [15] | DIPP | 11 type 1 diabetes | 18 | HLA, sex, period of birth | No | Lower 11-Eicosenoic acid. 1-Monopalmith, Arachidonic acid, Glutamic acid, L-5-Oxoproline, Linoleic acid, Oleic acid, Palmitic acid, Stearic acid. | 3 Months (plasma) |
| Sen, 2020 [17] | DIPP | 34 type 1 diabetes | 10 | No matching | No | 12 months: Increased BCAA1 and valine, Lower BCAA3, dihydroxyacetone phosphate, fructose&glucose-6-phosphate, Lysine, Myristic acid, Phenylalanine, phosphophoenolpyruvate and Tyrosine. | 12 months (non-fasting PBMC) |
| Li, 2020 [54] | TEDDY | 306 IA | 1-3 controls per case | 1:3 Clinical center, sex, type 1 diabetes family history. | HLA, risk SNPs | Lactulose, DHAA, Ethanolamine, Methanolphosphate (GADA-first); Lauric acid, Diglycerol, DHAA, γ-aminobutyric acid, Uric acid (IAA first) | 3 Months (plasma) |
| Johnson, 2019 [55] | TEDDY | 253 IA | 1-3 controls per case | 1:3 Clinical center, sex, type 1 diabetes family history, long distance protocol | HLA, age at blood draw | Dicarboxylic acids, Galactosylceramids, Phosphatodylethanolamines. | 9 Months (plasma) |
| Stanfill, 2019 [35] | TEDDY | 418 IA | 418 | 1:1 Clinical center, sex, type 1 diabetes family history. | n/a (conditional classification algorithm) | Influential metabolites: α-Tocopherol, Adipic acid, Glucose, Creatinine, Heptadecanoic acid, Hydroxybutanoic acid, Leucine, Isothreonic acid, Glyserol galactoside | First IA-positive sample (plasma) |
| Webb-Robertson, 2020 [56] | TEDDY | 157 IA | 157 | 1:1 Clinical center, sex, type 1 diabetes family history | n/a (machine learning) | Adipic and azelaic acid important features. Important features associated with lipid oxidation, phospholipase A2 signaling, and pentose phosphate pathway. | 6 months prior to IA |
| Frohnert 2020 [14] | DAISY | 20 IA | 25 | HLA, age, sex, FDR status | n/a (machine learning) | Top metabolic features: Vitamin C, 3-methyl-2-oxobutyrate, 4-hydroxyhippurate, pyroglutamine | Earliest sample prior to IA (9-15 months) compared to sample just prior to IA (serum). |
| Jørgenrud, 2017 [57] | MIDIA | 29 IA | 29 | 1:1 Birthdate, sex, county of residence | No | Tryptophan,3-hydroxybutiric acid, aspartic acid and cholesterol higher. Serine, Ornithine, Methionine and Tyrosine lower. | ~3 Months |
| Pflueger, 2011 [58] | BABYDIAB | 35 IA | 35 | 1:1 on Sex, age, birthdate, HLA | Parent with type 1 diabetes, HLA, fasting sample, age. | Lower methionine and hydroxyproline (but not pre-seroconversion) | At IA (serum). Difference in those that developed I ≤2 years of age |

Abbreviations: DIPP: Type 1 Diabetes Prediction and Prevention Study; TEDDY: The Environmental Determinants of Diabetes in the Young; DAISY: Diabetes Autoimmunity Study in the Young; MIDIA: Norwegian acronym of *Environmental causes of Type 1 Diabetes*; HLA: Human Leukocyte Antigen; SNP: Single nucleotide polymorphism, IA: Islet Autoimmunity; FDR: First Degree Relative; n/a: not applicable; PBMC: Peripheral blood mononuclear cells; DHAA: dehydroascorbic acid.

* Based on cases diagnosed or followed at Meyer Children's Hospital, Florence, Italy. Excluded children with neonatal or monogenic diabetes, low weight (<1800 g) pre-term newborns, newborns on parenteral nutrition and newborns receiving an exsanguination transfusion before the newborn screening test.

DIPP includes Finnish children at increased HLA genetic risk for type 1 diabetes (HLA-DQ2/DQ8 heterozygous, or DQ8/X, where X is anything but a type 1 diabetes-protective allele). Studies from Lamichhane and Sen only includes children from Tampere. TEDDY includes children at increased HLA genetic risk from the US, Finland, Sweden and Germany. DAISY includes children with HLA alleles conferring increased susceptibility for HLA and first-degree relatives of patients with type 1 diabetes. MIDIA includes Norwegian children carrying the highest HLA genetic risk for type 1 diabetes (HLA-DQ2/DQ8 heterozygous). BABYDIAB recruited German children born to mothers or fathers with type 1 diabetes.

### ESM Figure 1: Metabolite distributions in cases and controls

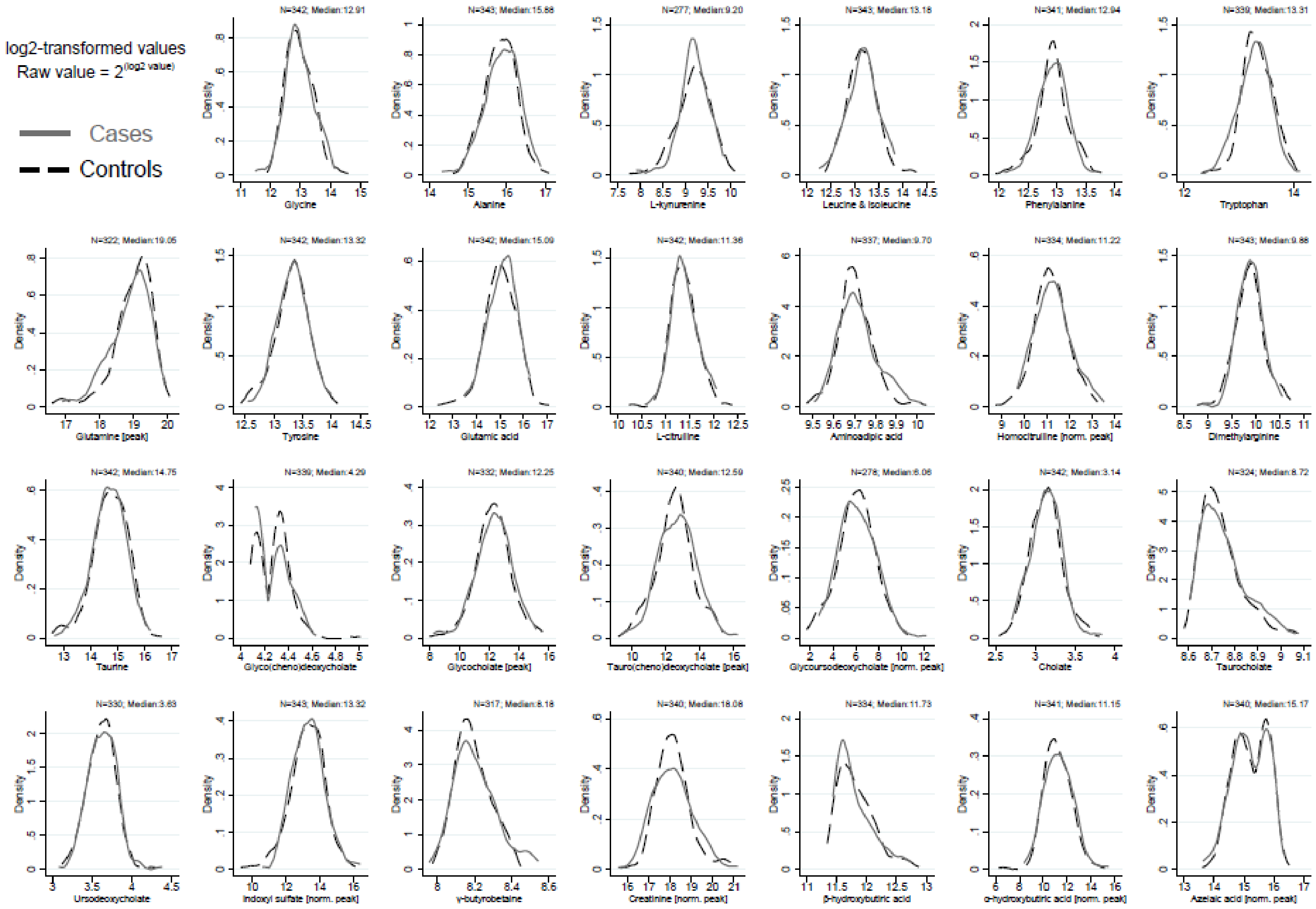

ESM Figure 1 shows the distribution of log2-transformed levels of the metabolites studied in random controls (dashed black line) and case children (solid grey line).

### ESM Figure 2:  Correlation matrix of metabolites

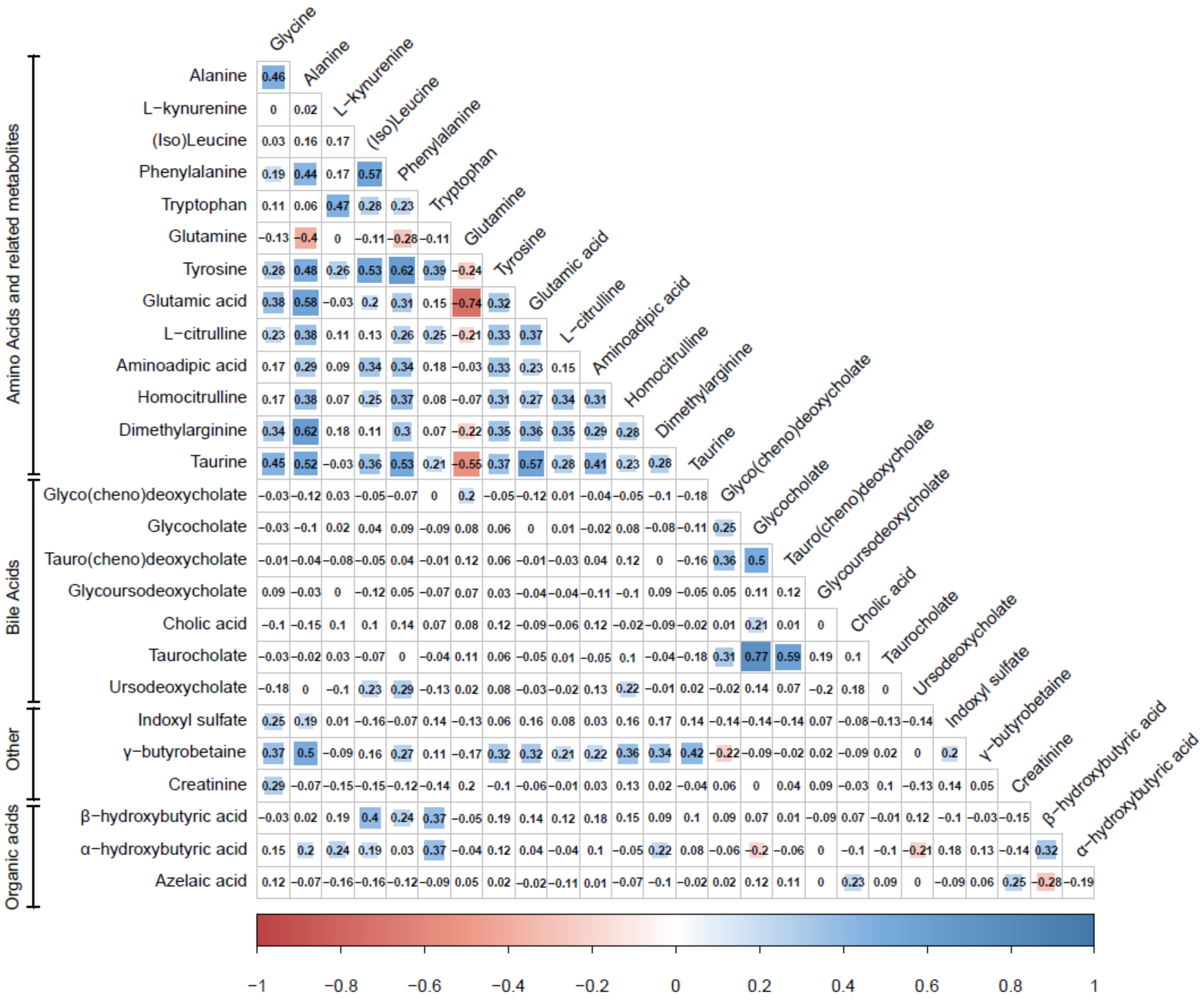

ESM Figure 2 shows the spearman correlation (Rho) between metabolites, as measured in the random controls. Significant (p≤0.01) correlations are marked with a coloured square, with size corresponding to the strength of correlation. Negative correlations are shown in red, while positive correlations are shown in blue. The figure was generated with R(version 3.6.1),[59] package *corrplot*.[60]

### ESM Figure 3: Categorical analysis

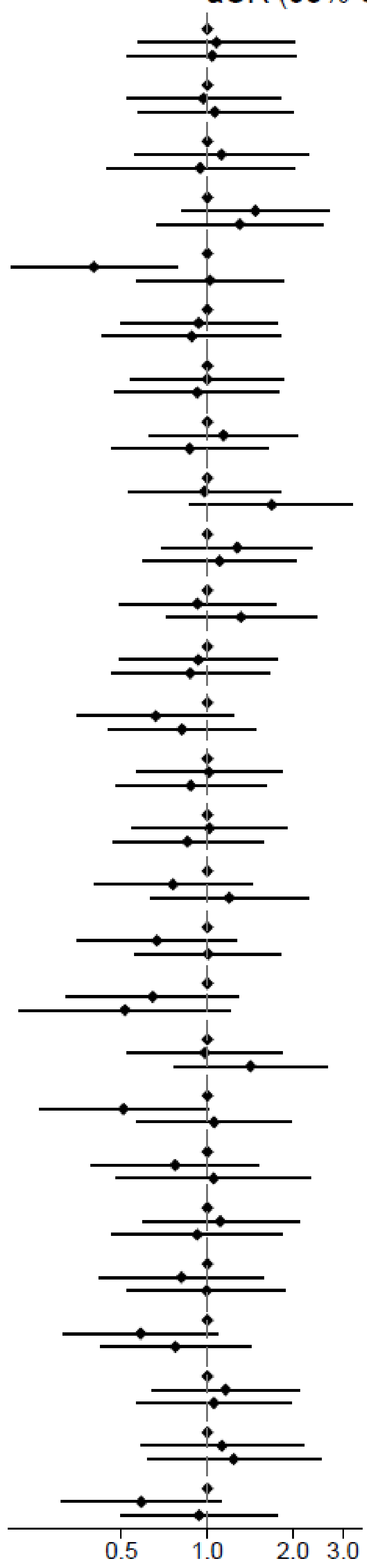

| Metabolite | Tertile | aOR (95% CI) |
|---|---|---|
| Glycine | 1st tertile | Ref. |
| | 2nd tertile | 1.08 (0.57 - 2.02) |
| | 3rd tertile | 1.04 (0.53 - 2.06) |
| Alanine | 1st tertile | Ref. |
| | 2nd tertile | 0.97 (0.52 - 1.80) |
| | 3rd tertile | 1.06 (0.57 - 1.99) |
| L-kynurenine | 1st tertile | Ref. |
| | 2nd tertile | 1.12 (0.56 - 2.26) |
| | 3rd tertile | 0.94 (0.44 - 2.02) |
| Leucine & Isoleucine | 1st tertile | Ref. |
| | 2nd tertile | 1.47 (0.81 - 2.68) |
| | 3rd tertile | 1.30 (0.66 - 2.54) |
| Phenylalanine | 1st tertile | Ref. |
| | 2nd tertile | 0.40 (0.20 - 0.78) |
| | 3rd tertile | 1.02 (0.57 - 1.85) |
| Tryptophan | 1st tertile | Ref. |
| | 2nd tertile | 0.93 (0.49 - 1.76) |
| | 3rd tertile | 0.88 (0.43 - 1.82) |
| Glutamine | 1st tertile | Ref. |
| | 2nd tertile | 1.00 (0.54 - 1.86) |
| | 3rd tertile | 0.92 (0.47 - 1.79) |
| Tyrosine | 1st tertile | Ref. |
| | 2nd tertile | 1.14 (0.62 - 2.08) |
| | 3rd tertile | 0.87 (0.46 - 1.63) |
| Glutamic acid | 1st tertile | Ref. |
| | 2nd tertile | 0.98 (0.53 - 1.80) |
| | 3rd tertile | 1.68 (0.87 - 3.25) |
| L-citrulline | 1st tertile | Ref. |
| | 2nd tertile | 1.27 (0.69 - 2.33) |
| | 3rd tertile | 1.10 (0.60 - 2.05) |
| Aminoadipic acid | 1st tertile | Ref. |
| | 2nd tertile | 0.92 (0.49 - 1.75) |
| | 3rd tertile | 1.31 (0.72 - 2.41) |
| Homocitrulline | 1st tertile | Ref. |
| | 2nd tertile | 0.93 (0.49 - 1.77) |
| | 3rd tertile | 0.87 (0.46 - 1.65) |
| Dimethylarginine | 1st tertile | Ref. |
| | 2nd tertile | 0.66 (0.35 - 1.24) |
| | 3rd tertile | 0.81 (0.45 - 1.48) |
| Taurine | 1st tertile | Ref. |
| | 2nd tertile | 1.01 (0.56 - 1.82) |
| | 3rd tertile | 0.88 (0.48 - 1.61) |
| Glyco(cheno)cholate | 1st tertile | Ref. |
| | 2nd tertile | 1.02 (0.55 - 1.89) |
| | 3rd tertile | 0.85 (0.46 - 1.56) |
| Glycocholic acid | 1st tertile | Ref. |
| | 2nd tertile | 0.76 (0.40 - 1.43) |
| | 3rd tertile | 1.19 (0.63 - 2.25) |
| Tauro(cheno)cholate | 1st tertile | Ref. |
| | 2nd tertile | 0.67 (0.35 - 1.27) |
| | 3rd tertile | 1.01 (0.56 - 1.82) |
| Glycoursocholate | 1st tertile | Ref. |
| | 2nd tertile | 0.64 (0.32 - 1.29) |
| | 3rd tertile | 0.51 (0.22 - 1.21) |
| Cholic acid | 1st tertile | Ref. |
| | 2nd tertile | 0.98 (0.52 - 1.84) |
| | 3rd tertile | 1.42 (0.76 - 2.64) |
| Taurocholic acid | 1st tertile | Ref. |
| | 2nd tertile | 0.51 (0.26 - 1.01) |
| | 3rd tertile | 1.06 (0.56 - 1.98) |
| Ursocholate | 1st tertile | Ref. |
| | 2nd tertile | 0.77 (0.39 - 1.52) |
| | 3rd tertile | 1.05 (0.48 - 2.31) |
| Indoxyl sulfate | 1st tertile | Ref. |
| | 2nd tertile | 1.11 (0.59 - 2.09) |
| | 3rd tertile | 0.92 (0.46 - 1.83) |
| γ-butyrobetaine | 1st tertile | Ref. |
| | 2nd tertile | 0.81 (0.42 - 1.58) |
| | 3rd tertile | 0.99 (0.52 - 1.89) |
| Creatinine | 1st tertile | Ref. |
| | 2nd tertile | 0.58 (0.31 - 1.09) |
| | 3rd tertile | 0.77 (0.42 - 1.42) |
| β-hydroxybutyric acid | 1st tertile | Ref. |
| | 2nd tertile | 1.16 (0.64 - 2.09) |
| | 3rd tertile | 1.05 (0.56 - 1.97) |
| α-hydroxybutyric acid | 1st tertile | Ref. |
| | 2nd tertile | 1.13 (0.58 - 2.17) |
| | 3rd tertile | 1.24 (0.61 - 2.50) |
| Azelaic acid | 1st tertile | Ref. |
| | 2nd tertile | 0.59 (0.31 - 1.12) |
| | 3rd tertile | 0.94 (0.50 - 1.75) |

ESM Figure 3 shows the results of a categorical analysis, with metabolites divided into tertiles. The lowest tertile is set as the reference (OR=1), and estimates are adjusted as in the main analysis. No metabolite showed significant deviation from linearity, except phenylalanine.

### ESM Figure 4: Metabolite Set Enrichment Analysis

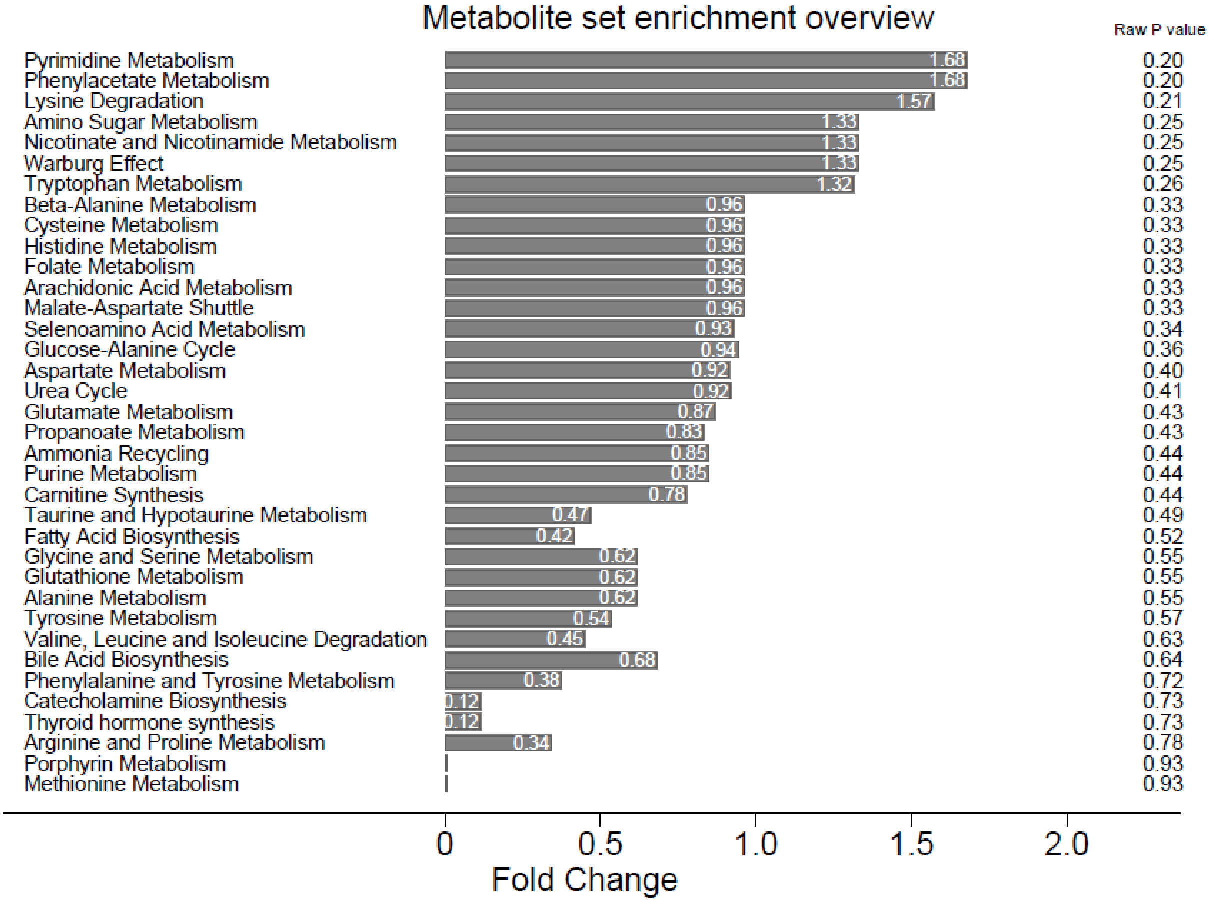

ESM Figure 4 shows the results from the metabolite set enrichment analysis. No metabolite set was significantly associated with case status.

#### ESM Figure 5: Scree plot of principal components and their association with later type 1 diabetes.

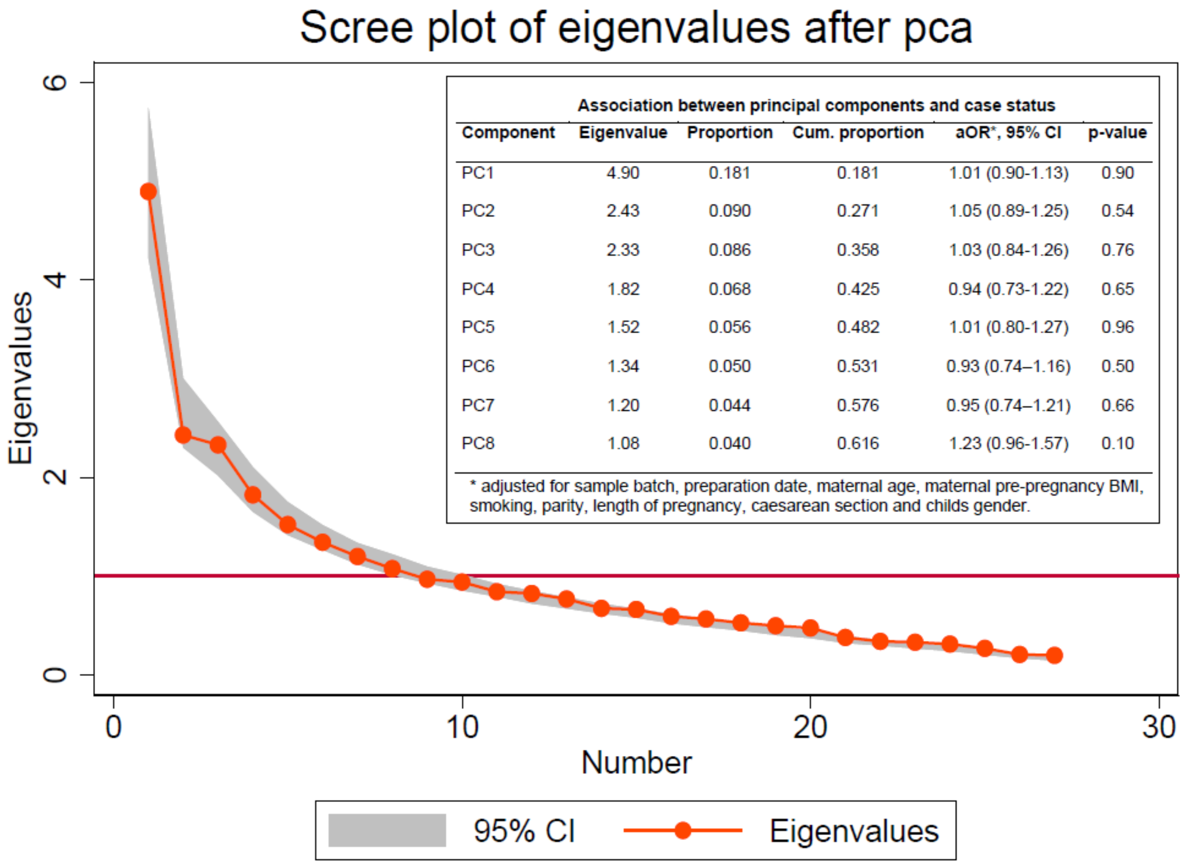

**Association between principal components and case status**

| Component | Eigenvalue | Proportion | Cum. proportion | aOR*, 95% CI | p-value |
|---|---|---|---|---|---|
| PC1 | 4.90 | 0.181 | 0.181 | 1.01 (0.90-1.13) | 0.90 |
| PC2 | 2.43 | 0.090 | 0.271 | 1.05 (0.89-1.25) | 0.54 |
| PC3 | 2.33 | 0.086 | 0.358 | 1.03 (0.84-1.26) | 0.76 |
| PC4 | 1.82 | 0.068 | 0.425 | 0.94 (0.73-1.22) | 0.65 |
| PC5 | 1.52 | 0.056 | 0.482 | 1.01 (0.80-1.27) | 0.96 |
| PC6 | 1.34 | 0.050 | 0.531 | 0.93 (0.74–1.16) | 0.50 |
| PC7 | 1.20 | 0.044 | 0.576 | 0.95 (0.74–1.21) | 0.66 |
| PC8 | 1.08 | 0.040 | 0.616 | 1.23 (0.96-1.57) | 0.10 |

* adjusted for sample batch, preparation date, maternal age, maternal pre-pregnancy BMI, smoking, parity, length of pregnancy, caesarean section and childs gender.

ESM Figure 5 plots the principal components in a scree plot (which plots the eigenvalues of component in descending order from largest to smallest), with eight components being above 1. These components were retained and used in a principal component analysis, with the inset table showing the association between these and later type 1 diabetes, their explained proportion and cumulative proportion. Proportion is how much of the overall variance that is explained by the component, and cumulative proportion is how much variance that is explained including all previous components. Plotting the principal components did not show any clear separation by case status (an example is show in ESM Figure 6.

#### ESM Figure 6: Principal Component plot

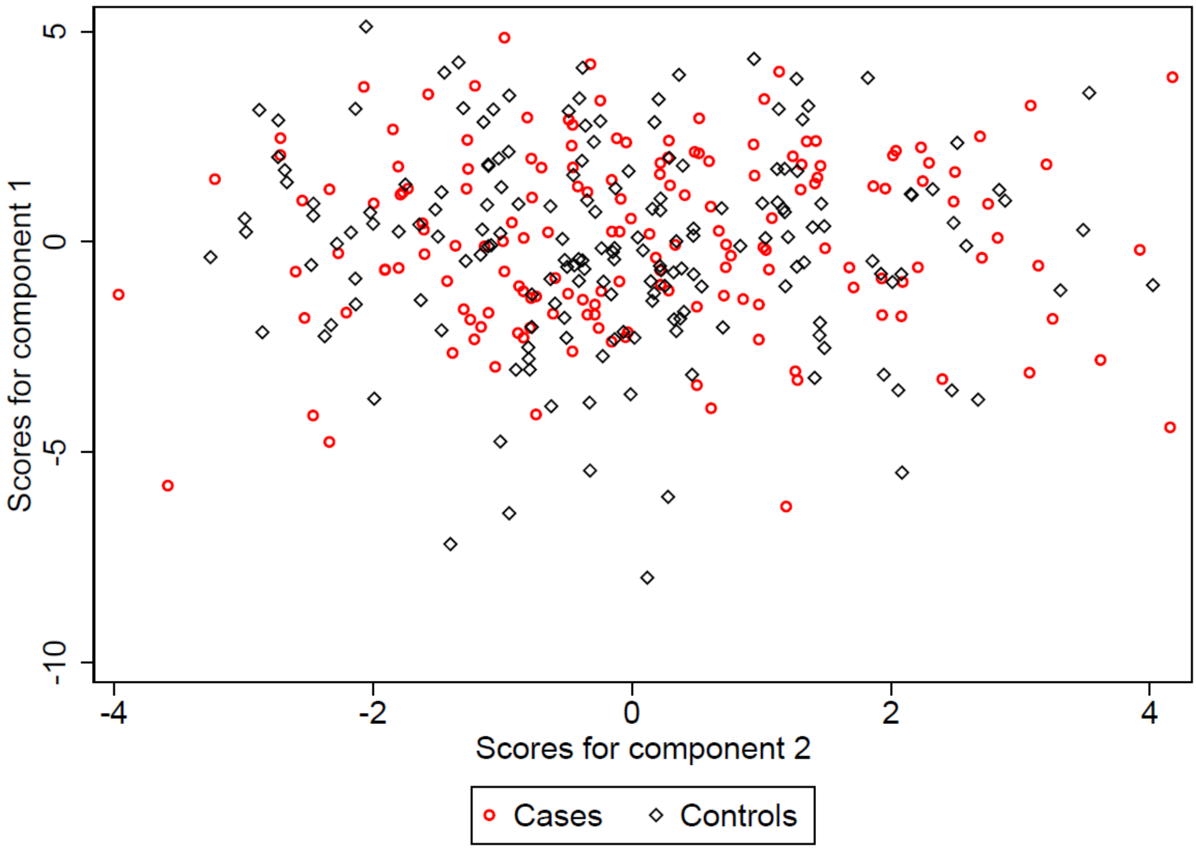

ESM Figure 6 shows the score of principal component (PC) 1 and PC2, with cases marked as hollow red circles and controls as hollow black diamonds. No other pairwise plotting of PCs showed separation.

### Electronic Supplementary References

46. Cooper, J.D., et al., *Meta-analysis of genome-wide association study data identifies additional type 1 diabetes risk loci.* Nat Genet, 2008. **40**(12): p. 1399-401.
47. Huang, J., et al., *1000 Genomes-based imputation identifies novel and refined associations for the Wellcome Trust Case Control Consortium phase 1 Data.* Eur J Hum Genet, 2012. **20**(7): p. 801-5.
48. Wellcome Trust Case Control, C., *Genome-wide association study of 14,000 cases of seven common diseases and 3,000 shared controls.* Nature, 2007. **447**(7145): p. 661-78.
49. Todd, J.A., et al., *Robust associations of four new chromosome regions from genome-wide analyses of type 1 diabetes.* Nat Genet, 2007. **39**(7): p. 857-64.
50. Heinig, M., et al., *A trans-acting locus regulates an anti-viral expression network and type 1 diabetes risk.* Nature, 2010. **467**(7314): p. 460-4.
51. Wallace, C., et al., *The imprinted DLK1-MEG3 gene region on chromosome 14q32.2 alters susceptibility to type 1 diabetes.* Nat Genet, 2010. **42**(1): p. 68-71.
52. Smyth, D.J., et al., *FUT2 nonsecretor status links type 1 diabetes susceptibility and resistance to infection.* Diabetes, 2011. **60**(11): p. 3081-4.
53. Vistnes, M., et al., *Plasma immunological markers in pregnancy and cord blood: A possible link between macrophage chemo-attractants and risk of childhood type 1 diabetes.* Am J Reprod Immunol, 2018. **79**(3).
54. Li, Q., et al., *Longitudinal Metabolome-Wide Signals Prior to the Appearance of a First Islet Autoantibody in Children Participating in the TEDDY Study.* Diabetes, 2020. **69**(3): p. 465-476.
55. Johnson, R.K., et al., *Metabolite-related dietary patterns and the development of islet autoimmunity.* Sci Rep, 2019. **9**(1): p. 14819.
56. Webb-Robertson, B.M., et al., *Prediction of the development of islet autoantibodies through integration of environmental, genetic, and metabolic markers.* J Diabetes, 2020.
57. Jørgenrud, B., et al., *Longitudinal plasma metabolic profiles, infant feeding, and islet autoimmunity in the MIDIA study.* Pediatr Diabetes, 2017. **18**(2): p. 111-119.
58. Pflueger, M., et al., *Age- and islet autoimmunity-associated differences in amino acid and lipid metabolites in children at risk for type 1 diabetes.* Diabetes, 2011. **60**(11): p. 2740-7.
59. R Core Team, *R: A language and environment for statistical computing*. 2019, R foundation for statistical computing: Vienna, Austria.
60. Wei, T. and V. Simko, *R package "corrplot": Visualization of a Correlation Matrix*. 2017.
